# External Evaluation and Calibration Drift of Explainable Machine Learning Models for Acute and Chronic GVHD Prediction After Allogeneic HSCT

**DOI:** 10.64898/2026.06.14.26355639

**Authors:** Naveed Syed, Nausheen Ahmed, Mohamed Abuhaleeqa, Fatema Mohammed Al Kaabi, Ahmad Raza, Ajlan Al Zaki, Fatin Sammour, Yaser Alkhatib, Dharmesh Gopalakrishnan, Imrana Afrooz, Moussab Damlaj, Husam Abu Jazar, Hikmat Abdel-Razeq, Khalid Halahleh, Mohammad Yaqub, Shahrukh Hashmi

## Abstract

**Objectives:** Machine learning models for graft-versus-host disease (GVHD) after allogeneic hematopoietic stem cell transplantation (allo-HSCT) are rarely evaluated externally or for calibration. We developed explainable models for grade II–IV acute and moderate-to-severe chronic GVHD and evaluated their transportability across multiple cohorts.

**Materials and methods:** Gradient boosting models were trained on three CIBMTR registry-derived datasets (N = 2,509) and externally evaluated in six cohorts (N = 14,788), including a Middle Eastern cohort. We assessed discrimination (AUROC), calibration (logistic recalibration intercept and slope; Brier score), and SHAP feature explanations.

**Results:** Internal discrimination was higher for chronic (AUROC 0.736) than acute GVHD (0.624); chronic calibration was good (intercept +0.258, slope 1.073), whereas the acute model was underdispersed (slope 1.802). Externally, the chronic model retained modest, above-chance discrimination (AUROC 0.54–0.64), whereas acute performance fell toward chance (0.52–0.57). Calibration drifted unpredictably, overestimating risk in 9 of 10 evaluable cohort–outcome combinations (intercepts −0.78 to +0.15; slopes 0.12 to 1.98; Brier 0.23–0.27).

**Discussion:** Routine registry variables carry a transportable signal for chronic GVHD, but acute GVHD likely requires additional biological or clinical predictors. Neither the direction nor the magnitude of calibration drift could be anticipated from training performance.

**Conclusion:** Internal performance does not guarantee external utility. Cohort-specific recalibration and routine calibration reporting are required before any clinical use; this study offers a reusable template for multi-cohort external evaluation of clinical prediction models.

**Lay Summary:** After a stem cell transplant from a donor, the donor’s immune cells can attack the patient’s body, a complication called graft-versus-host disease (GVHD). It can appear early (acute) or months later (chronic), and knowing who is at highest risk before transplant could help doctors plan prevention and follow-up. We built computer models that estimate GVHD risk using information routinely recorded at transplant, trained them on 2,509 patients from a large US transplant registry, and then tested them on six other groups totalling almost 15,000 patients, including patients from the United Arab Emirates and Jordan. The model for chronic GVHD kept a modest ability to separate higher-from lower-risk patients in every group, but the model for acute GVHD performed little better than chance outside the data it was built on. In almost every new group, both models overestimated how many patients would develop GVHD, by amounts that could not be predicted in advance. This shows that a model that works well where it was developed may not work elsewhere, and that its risk estimates must be checked and adjusted locally before it is ever used to guide care.

## Introduction

Graft-versus-host disease (GVHD) remains a major complication following allogeneic hematopoietic stem cell transplantation (allo-HSCT), particularly in patients with acute myeloid leukemia (AML), acute lymphoblastic leukemia (ALL), and myelodysplastic syndromes (MDS), where both acute and chronic GVHD continue to adversely impact overall survival (OS) despite advances in transplant strategies^1,2,3,4^. Accurate prediction of GVHD risk remains challenging due to the complex interplay of immunological, donor-related, and transplant-specific factors^4,5^.

Existing GVHD prediction models demonstrate modest discrimination and limited external generalizability (C-statistics ~0.62–0.65 and AUROC values ~0.61–0.64) ^6,7,8^. Biomarker-based models achieve higher discrimination (~0.65–0.70) but are limited by restricted clinical applicability ^9, 10^. Similarly, while attempts using dynamic EHR vital signs have shown promise, they remain largely single-center and unvalidated, highlighting the need to benchmark static registry models against these emerging dynamic approaches.

Machine learning approaches can model nonlinear interactions among transplant-related variables, but most published models lack external evaluation or depend on highly granular inputs limiting routine applicability ^8,11,12^. External evaluation across heterogeneous cohorts remains a key unmet need ^13,14^.

To address these limitations, we conducted a multi-cohort evaluation of our registry-trained machine learning for GVHD prediction. Using explainable gradient boosting models built from routinely available clinical variables with support for HLA compatibility metrics from allele-level typing where available, we predicted acute and chronic GVHD in patients undergoing allo-HSCT for AML, ALL, and MDS, then externally evaluated these models across six external cohorts, explicitly characterizing discrimination and calibration drift under real-world feature missingness. Model behavior was interpreted using SHapley Additive exPlanations (SHAP). Our aim is to establish how such models should be validated before clinical use: a blueprint emphasizing external, multi-cohort evaluation and routine calibration reporting. The framework is intended for research and methodological evaluation, not clinical deployment, pending prospective evaluation.

## Methods

### Study Design and Data Sources

We developed and externally evaluated an explainable machine learning framework for acute and chronic GVHD following allo-HSCT, using outcomes as adjudicated in each originating registry. Acute GVHD was the grade II–IV endpoint (modified Glucksberg/consensus grading) within the 100-day window; chronic GVHD was the NIH moderate-to-severe endpoint. In the UAE+KHCC cohort these thresholds could not be confirmed, so it was excluded from standardized acute-GVHD calibration metrics but retained descriptively.

Nine datasets were used: three publicly released CIBMTR-derived datasets (combined N=2,509) for development and six external evaluation cohorts, restricted to AML, ALL, and MDS; source studies, eligibility, transplant periods, and outcome definitions are summarized in **Supplementary Document 15**. Training datasets were selected for availability of clinically relevant variables, while several external cohorts had substantial missingness, enabling assessment of generalizability under real-world constraints. The study followed TRIPOD-AI recommendations; **Figure 1** shows the study workflow.

**Figure 1.**
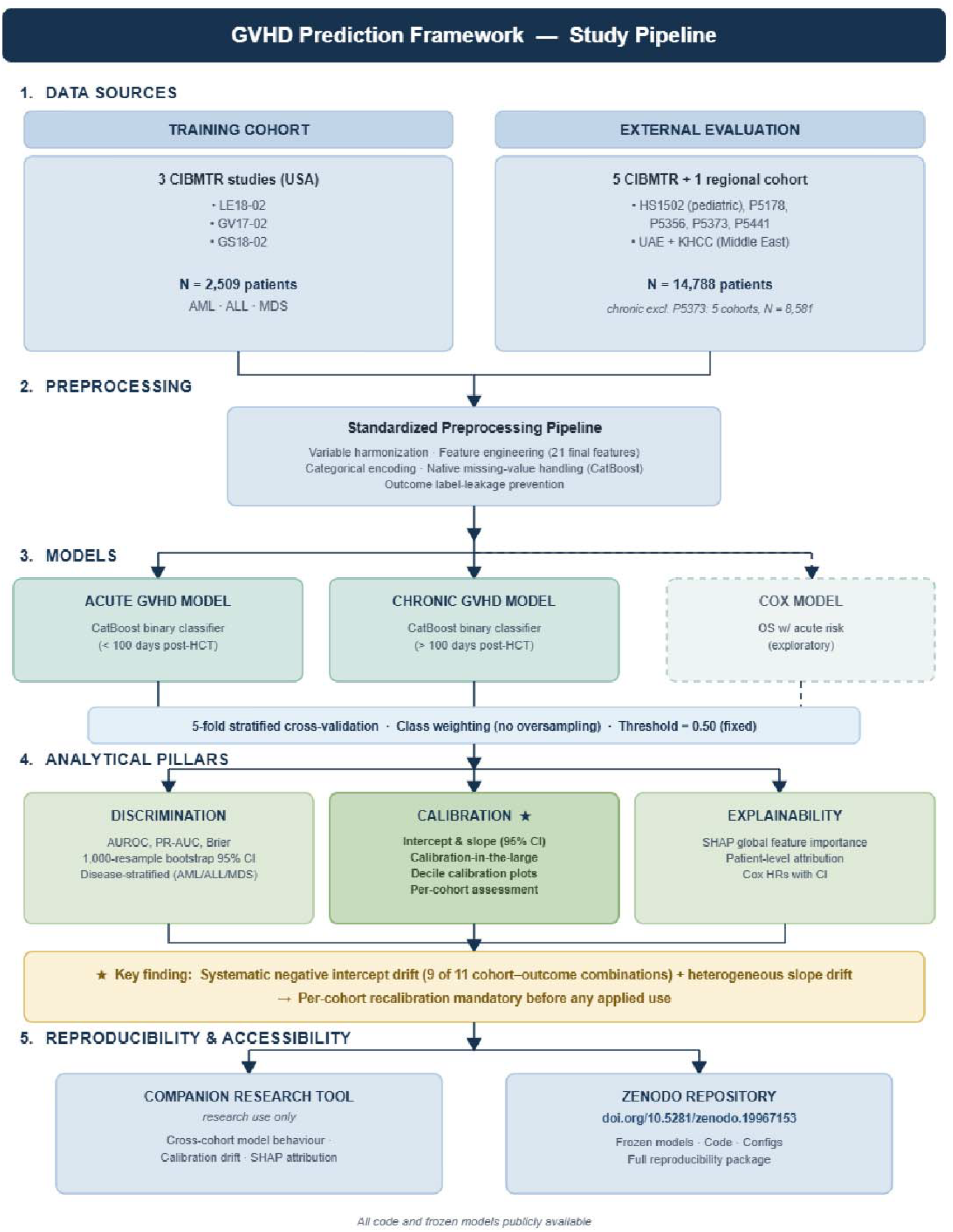
Schematic overview of the study workflow, preprocessing pipeline, modelling strategy, external evaluation design, and analytical framework.

The regional Middle Eastern cohort (UAE+KHCC) represents a distinct real-world paradigm: these patients underwent allo-HSCT abroad with regional follow-up and no integrated local registry, so strict grade II–IV acute-GVHD thresholds could not be harmonized with CIBMTR standards. It was therefore treated as an uncalibrated stress-test for registry-to-clinical-record endpoint mismatch.

### Preprocessing, Features, and Model Development

A dataset was eligible as an external cohort if it contained the GVHD outcome of interest and a minimum core feature set comprising recipient age, disease type (AML/ALL/MDS), donor type, graft source, conditioning intensity, GVHD prophylaxis, and HLA match category. Patients were retained regardless of missingness in non-core variables; no complete-case exclusion was performed, and missing values were handled natively by CatBoost. This design deliberately preserved real-world data incompleteness so that model behaviour could be assessed under the feature missingness typical of registry data.

### Data Preprocessing and Feature Engineering

A standardized pipeline harmonized naming, formatting, and missing-value handling; missing values were handled natively by CatBoost without imputation. The primary HLA variable was a four-category match ratio (full, partial, mismatched, unknown), a harmonized proxy rather than an allele-level count. Outcome-defining variables were excluded to prevent leakage. Gradient boosting (CatBoost) was used, with separate acute and chronic models on identical schemas and class weights inversely proportional to prevalence. Performance was optimized via stratified k-fold cross-validation with out-of-fold predictions, then frozen for external evaluation. From 39 candidate variables, feature selection combined clinical relevance, availability, and univariate signal; the final model included 21 features. The complete feature audit, including per-feature missingness, univariate AUC values, hyperparameter settings, and per-fold performance metrics are detailed in **Supplementary Documents 1** and **2**.

### Evaluation and Calibration

Internal evaluation used stratified cross-validation, with disease-specific analyses for AML, ALL, and MDS, and external evaluation across six external cohorts assessed overall and by subgroup. Threshold optimization gave optimal thresholds of 0.49 for acute (balanced accuracy 0.597) and 0.50 for chronic GVHD (0.680); a fixed 0.50 threshold was used for comparability, though prospective deployment should re-optimize locally. Calibration was assessed in the training cohort (out-of-fold, 5-fold) and all external cohorts by logistic recalibration: the intercept reflects systematic over-/under-estimation (negative = overestimation) and the slope over-(<1) or under-spread (>1), with 1 indicating ideal calibration^15^. 95% CIs came from analytical standard errors. Calibration-in-the-large, Brier scores, calibration plots, and bootstrap AUROC CIs (1,000 resamples) were also reported. Interpretability used SHAP for global and patient-level attribution.

### Reproducibility and Data Availability

The frozen package (GVHD Intel Pro v1.0) is publicly available at https://doi.org/10.5281/zenodo.19967153, enabling exact reconstruction of all analyses^16^. An open-access companion tool [https://app.gvhdriskcalculator.com/] is for research use only and is not a validated clinical calculator. Full details are in the Supplementary Methods. Analyses of CIBMTR-derived datasets followed registry data-use policies with no patient-identifiable information accessed; the UAE+KHCC cohort comprised retrospectively collected, de-identified allogeneic HCT data from institutions in the United Arab Emirates and Jordan, approved by the respective institutional review boards and ethics committees.

## Results

### Internal Validation

In 5-fold cross-validation, the acute GVHD model demonstrated a mean fold AUROC of 0.632 ± 0.023 with an aggregated out-of-fold (OOF) AUROC of 0.624; PR-AUC was 0.54 and Brier score 0.24 (training cohort N=2,509, 1,091 events, prevalence 0.43). The chronic GVHD model demonstrated a mean AUROC of 0.740 ± 0.024 with an aggregated OOF AUROC of 0.736; PR-AUC was 0.74 and Brier score 0.21 (1,426 events, prevalence 0.57). Disease-wise AUROC values for acute GVHD were 0.62 (AML), 0.62 (ALL), and 0.64 (MDS); for chronic GVHD, 0.74 (AML), 0.68 (ALL), and 0.79 (MDS). Performance metrics across cohorts are summarized in **Table 1** and **Table 2**; disease-wise ROC and SHAP plots are shown in **Figure 2**.

**Table 1.**
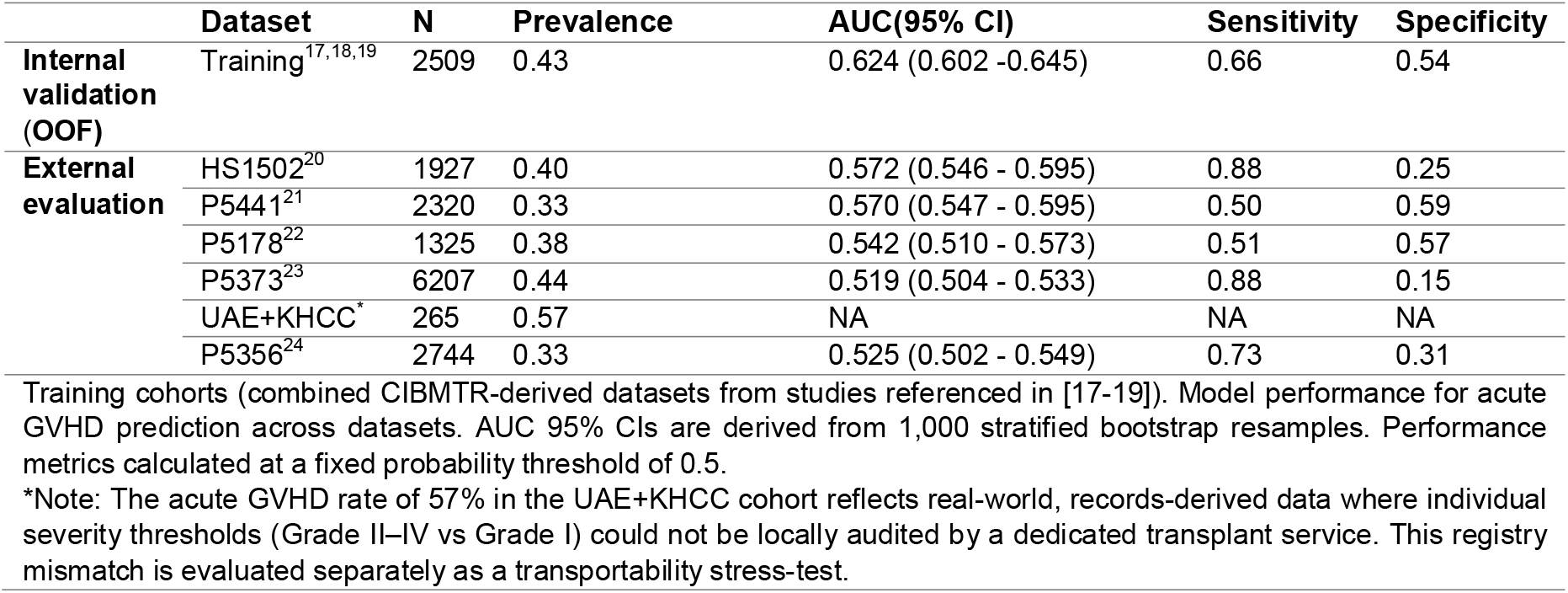
Model performance for acute GVHD prediction across datasets.

**Table 2.** Model performance for chronic GVHD prediction across datasets.

|  | Dataset | N | Prevalence | AUC (95% CI) | Sensitivity | Specificity |
| --- | --- | --- | --- | --- | --- | --- |
| **Internal Validation (OOF)** | Training<sup>17,18,19</sup> | 2509 | 0.57 | 0.736 (0.716 - 0.756) | 0.80 | 0.56 |
| **External evaluation** | P5356<sup>24</sup> | 2744 | 0.41 | 0.635 (0.614 - 0.656) | 0.11 | 0.95 |
|  | P5178<sup>22</sup> | 1325 | 0.29 | 0.627 (0.594 - 0.662) | 0.66 | 0.56 |
|  | UAE+KHCC | 265 | 0.42 | 0.542 (0.465 - 0.615) | 0.72 | 0.33 |
|  | P5441<sup>21</sup> | 2320 | 0.30 | 0.612 (0.590 - 0.637) | 0.44 | 0.71 |
|  | HS1502<sup>20</sup> | 1927 | 0.31 | 0.565 (0.536 - 0.593) | 0.50 | 0.63 |

Training cohorts (combined CIBMTR-derived datasets from studies referenced in [17-19]). Model performance for chronic GVHD prediction across datasets. AUC 95% CIs are derived from 1,000 stratified bootstrap resamples. Performance metrics calculated at a fixed probability threshold of 0.5. P5373 was excluded from chronic GVHD evaluation because the public-release dataset did not include a chronic GVHD outcome variable; chronic evaluation therefore comprises 5 external cohorts (n = 8,581 patients combined).

**Figure 2.**
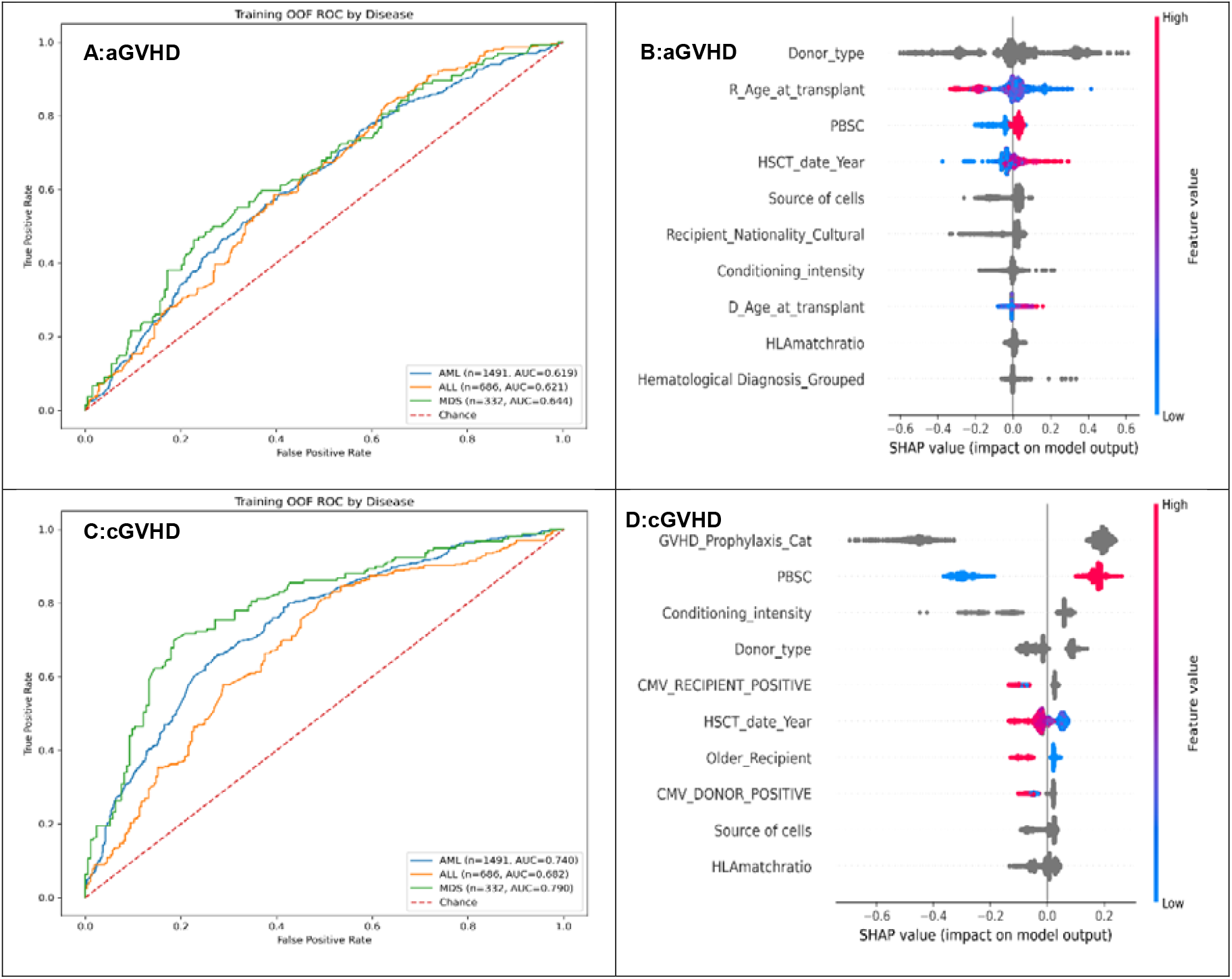
Training performance and feature importance of GVHD prediction models. Internal (out-of-fold) training performance and feature importance of the GVHD prediction models. (A– B) Acute GVHD; (C–D) Chronic GVHD. Disease-stratified ROC curves (A, C) show out-of-fold discrimination in the training cohort (acute: AML 0.62, ALL 0.62, MDS 0.64; chronic: AML 0.74, ALL 0.68, MDS 0.79), highest in MDS for both outcomes. SHAP summary plots (B, D) show the top 10 predictors and the direction of their effect on model output; leading contributors were donor type, recipient age, and graft source (PBSC) for acute GVHD, and GVHD prophylaxis, PBSC use, conditioning intensity, and donor type for chronic GVHD. The dashed diagonal denotes chance performance.

### External Evaluation

External evaluation spanned six external cohorts (N = 14,788; cohort sizes 265– 6,207). For acute GVHD, AUROC ranged from 0.52 to 0.57 (**Table 1**); for chronic GVHD, from 0.54 to 0.64 (**Table 2)**. Disease-wise values below are from the best-performing cohorts (acute, P5441; chronic, P5356): acute AUROC 0.55 (AML), 0.59 (ALL), 0.60 (MDS); chronic 0.63 (AML), 0.71 (ALL), 0.64 (MDS). **Figure 3** summarizes AUROC alongside feature availability across cohorts, and **Figure 4** shows representative subgroup ROC and SHAP plots.

**Figure 3.**
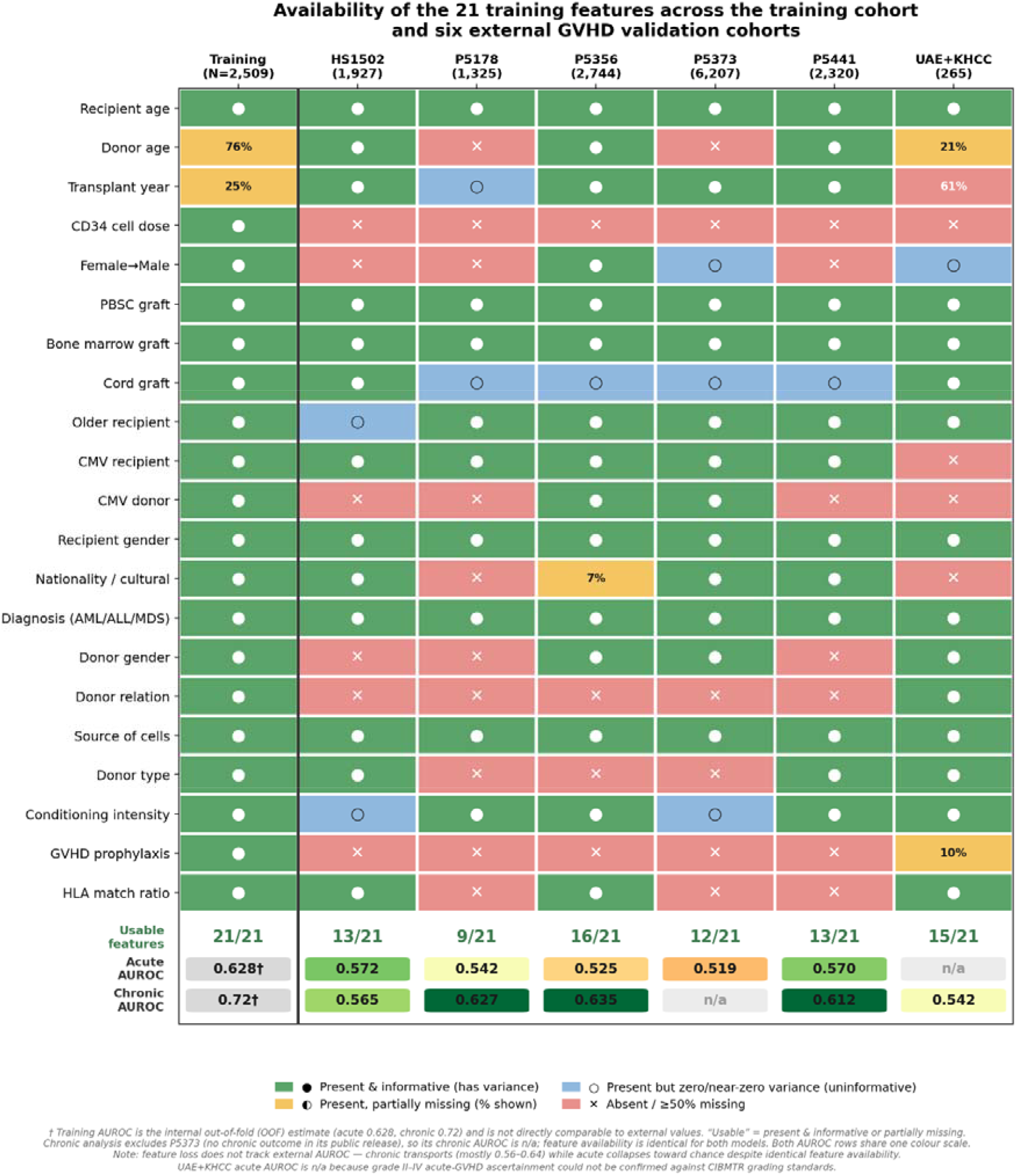
Availability of the 21 model features across the training cohort and the six external evaluation cohorts, shown alongside external discrimination. Each cell indicates whether a feature was present and informative, present but partially missing, present but uninformative (zero/near-zero variance), or absent/≥50% missing; the lower strips give the number of usable features and the external AUROC for acute and chronic GVHD (training values are internal out-of-fold estimates; chronic excludes P5373, which lacks a chronic outcome). Feature availability is a property of the source dataset and is therefore identical for the acute and chronic models. Despite this shared availability, chronic discrimination remained moderate (AUROC ≈ 0.54–0.64) while acute discrimination collapsed toward chance (≈ 0.52–0.57) across the same cohorts, indicating that reduced feature availability in the external cohorts likely contributed to the overall external decline.

**Figure 4.**
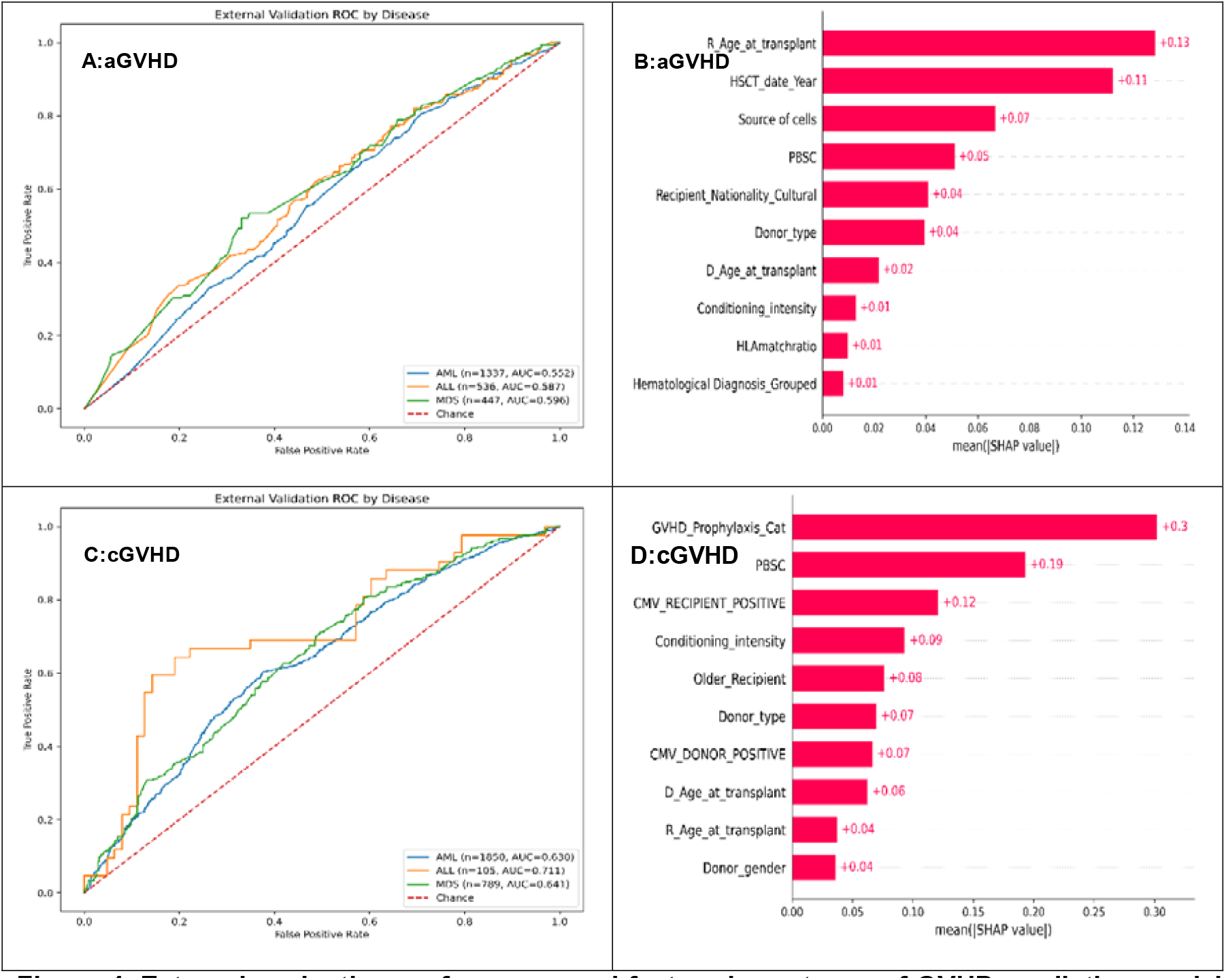
External evaluation performance and feature importance of GVHD prediction models in the best-performing cohorts. External evaluation performance and feature importance in the best-performing external cohorts (best case, not representative of typical external performance). (A–B) Acute GVHD, P5441 cohort; (C–D) Chronic GVHD, P5356 cohort. Disease-stratified ROC curves (A, C) show the highest external discrimination observed (acute P5441: AML 0.55, ALL 0.59, MDS 0.60; chronic P5356: AML 0.63, ALL 0.71, MDS 0.64). SHAP summary plots (B, D) show the most influential predictors, recipient age, transplant year, and graft source for acute GVHD, and GVHD prophylaxis, PBSC use, and CMV status for chronic GVHD. The dashed diagonal denotes chance performance.

Detailed disease-wise AUROC analyses, feature missingness profiles, subgroup ROC curves, cohort-specific calibration plots, and SHAP feature importance plots are provided separately for each external evaluation dataset for both acute and chronic GVHD in **Supplementary Documents 3-14**.

## Calibration

Calibration intercept, slope, calibration-in-the-large (CITL), and Brier score were computed across the training cohort (out-of-fold predictions) and across all external evaluation cohorts with available outcome data (five cohorts for acute GVHD; five cohorts for chronic GVHD, with P5373 excluded due to absence of chronic GVHD outcome data). Detailed calibration metrics are presented in **Table 3**, with calibration plots for best calibration cohorts shown in **Figure 5**.

**Table 3.** Calibration metrics for the GVHD prediction models across all cohorts.

| Target | Cohort | N | Events | Prev | Intercept (95% CI) | Slope (95% CI) | CITL | Brier |
| --- | --- | --- | --- | --- | --- | --- | --- | --- |
| **Acute** | Training (OOF) | 2,509 | 1,091 | 0.43 | -0.274 (-0.355 to -0.193) | 1.802 (1.455 to 2.149) | +0.01 | 0.241 |
|  | HS1502 | 1,927 | 764 | 0.40 | -0.777 (-0.923 to -0.630) | 1.982 (1.374 to 2.589) | -0.146 | 0.256 |
|  | P5178 | 1,325 | 502 | 0.38 | -0.451 (-0.572 to -0.330) | 0.207 (-0.028 to 0.442) | -0.070 | 0.248 |
|  | P5356 | 2,744 | 906 | 0.33 | -0.720 (-0.801 to -0.638) | 0.121 (-0.055 to 0.298) | -0.193 | 0.268 |
|  | P5373 | 6,207 | 2,744 | 0.44 | -0.300 (-0.376 to -0.225) | 0.430 (0.073 to 0.788) | -0.097 | 0.256 |
|  | P5441 | 2,320 | 769 | 0.33 | -0.720 (-0.808 to -0.633) | 0.913 (0.571 to 1.255) | -0.171 | 0.248 |
| **Chronic** | Training (OOF) | 2,509 | 1,426 | 0.57 | +0.258 (+0.172 to +0.344) | 1.073 (0.955 to 1.192) | +0.07 | 0.210 |
|  | HS1502 | 1,927 | 598 | 0.31 | -0.748 (-0.847 to -0.648) | 0.831 (0.384 to 1.278) | -0.173 | 0.243 |
|  | P5178 | 1,325 | 386 | 0.29 | -0.777 (-0.900 to -0.655) | 1.381 (0.999 to 1.763) | -0.181 | 0.231 |
|  | P5356 | 2,744 | 1,125 | 0.41 | +0.154 (+0.040 to +0.267) | 1.223 (1.016 to 1.430) | +0.015 | 0.230 |
|  | P5441 | 2,320 | 691 | 0.30 | -0.628 (-0.730 to -0.526) | 1.342 (1.019 to 1.666) | -0.155 | 0.227 |
|  | UAE+KHCC | 265 | 112 | 0.42 | -0.331 (-0.579 to -0.084) | 0.366 (-0.320 to 1.051) | -0.090 | 0.254 |

Calibration intercept and slope were estimated by logistic recalibration of out-of-fold (training) or out-of-cohort (external) predicted probabilities, with 95% CIs from analytical standard errors. Calibration-in-the-large (CITL) is the difference between the mean observed event rate and the mean predicted probability; negative values indicate the model overestimates risk. An ideal calibration slope equals 1.0; slopes below 1.0 indicate over-spread (overconfident) predictions and slopes above 1.0 indicate under-spread (underconfident) predictions; a slope 95% CI that does not include 1.0 indicates statistically significant miscalibration in spread. All external cohorts except P5356 chronic showed negative intercepts, indicating systematic risk overestimation. P5373 contains no chronic GVHD outcome and is therefore absent from the chronic rows. UAE+KHCC is excluded from the acute rows because grade II–IV acute-GVHD ascertainment could not be confirmed; its acute results are reported descriptively in the text. Prev = prevalence; OOF = out-of-fold; CITL = calibration-in-the-large.

**Figure 5.**
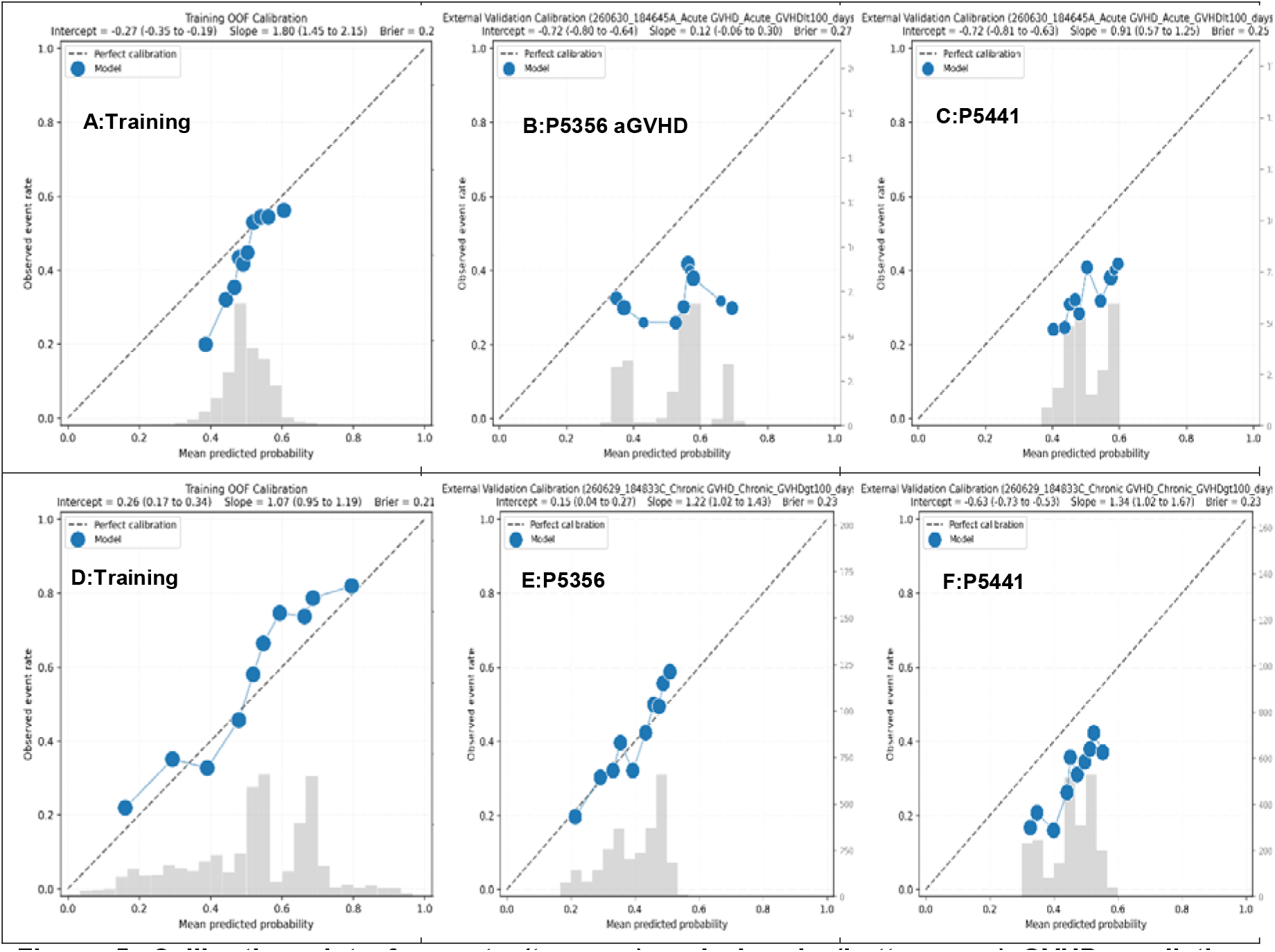
Calibration plots for acute (top row) and chronic (bottom row) GVHD prediction models across training and best calibration external evaluation cohorts. (A & D training cohorts, B & E P5356 cohort, C & F P5441cohorts) Filled circles represent observed event rates within deciles of predicted probability; dashed diagonal indicates ideal calibration. Calibration intercept and slope (95% CI) are shown in each panel title. Histogram overlays represent predicted probability distributions.

In the training cohort, the acute GVHD model had a calibration intercept of −0.274 (95% CI −0.355 to −0.193) and slope 1.802 (1.455 to 2.149); the chronic GVHD model had intercept +0.258 (+0.172 to +0.344) and slope 1.073 (0.955 to 1.192). Brier scores were 0.240 (acute) and 0.210 (Chronic).

Across external cohorts, calibration drift was substantial and heterogeneous. Negative intercept drift occurred in 9 of 10 evaluable external cohort–target combinations (intercepts −0.30 to −0.78); the only exception was P5356 chronic (+0.15, +0.040 to +0.267). Slope 95% CIs excluded 1.0 in four acute combinations : HS1502 (1.98, 1.37 to 2.59), P5178 (0.21, −0.03 to 0.44), P5356 (0.12, −0.06 to 0.30), P5373 (0.43, 0.07 to 0.79), and two chronic : P5356 (1.22, 1.02 to 1.43) and P5441 (1.34, 1.02 to 1.67). CITL ranged from −0.19 to +0.015, smallest for P5356 chronic (+0.015).

### Real-World Transportability and Endpoint Stress-Testing (UAE+KHCC)

In the regional Middle Eastern cohort (UAE+KHCC, N=265), medical-record documentation could not be mapped to strict CIBMTR grade II–IV criteria, so the cohort was excluded from standardized acute-GVHD discrimination and calibration analyses (Tables 1 and 3). Descriptively, apparent acute-GVHD prevalence was higher than in CIBMTR-derived cohorts (56.6% vs 33–44%), and the acute calibration slope was negative (−0.31, 95% CI −1.21 to 0.60) - consistent with loss or reversal of risk ordering from endpoint-ascertainment mismatch rather than conventional miscalibration. Chronic GVHD, a binary diagnosis documented by transplant physicians and less sensitive to retrospective grading than acute staging, was retained (AUROC 0.542; **Table 2**). Where endpoints cannot be harmonized, calibration fails informatively; such cohorts should be reported descriptively, not pooled into transportability conclusions.

## Discussion

We evaluated our registry-trained machine learning for GVHD prediction, redeveloping our proof-of-concept framework into explainable acute- and chronic-GVHD models for allo-HSCT patients with AML, ALL, and MDS, and testing transportability across six external cohorts^25^. Internal discrimination was moderate for chronic and modest for acute GVHD (AUROC 0.74 and 0.62); externally, the chronic model held modest discrimination (0.54–0.64) across all cohorts, whereas acute fell to 0.52–0.57, near chance in most (**Table 1 & Table 2**). The drop is the central finding, but it is not uniform: chronic GVHD kept a valid, if weaker, ranking, so its external failure was mainly calibration drift, whereas acute discrimination largely collapsed.

This discrimination matches prior registry-trained models, which typically report C-statistics of 0.60–0.65 and are rarely externally validated^8,11^; Lee et al. reported 0.618 for acute GVHD and 0.640 for a composite endpoint^26^, and Tang et al. 0.659 using longitudinal EHR data^11^, whereas our framework relies only on pre-transplant and transplant-related variables. Benchmarked against the CIBMTR acute-GVHD risk index^27^, our internal acute AUROC (0.62) matched that score’s validation discrimination (0.615) but was lower in the two fully adult external cohorts (P5441 0.57, P5178 0.54). The rest were unsuitable for comparison: P5356 and P5373 mix adult and pediatric recipients, HS1502 is pediatric, and UAE+KHCC is records-derived with uncertain grading, leaving a patient-level head-to-head as a priority for prospective evaluation. That even a purpose-built adult score reaches only 0.615 points to a low ceiling on the acute-GVHD signal in routine pre-transplant variables^28,12,14^, grade II–IV acute GVHD is intrinsically hard to predict from baseline registry data, and honest reporting of this limit beats an over-optimistic single-cohort estimate.

For moderate-to-severe chronic GVHD, discrimination was higher internally (0.74) and reached 0.61–0.64 in the larger external cohorts (P5356 0.635; P5178 0.627; P5441 0.612), indicating a more transportable baseline signal. No externally validated pre-transplant score exists for this endpoint, existing scores estimate survival or non-relapse mortality in patients who already have chronic GVHD or rely on post-transplant biomarkers or longitudinal records, so ours provides the first multi-cohort external assessment of baseline chronic-GVHD prediction^10^. Because this endpoint drives late morbidity and impaired quality of life, even modest discrimination is relevant for risk-stratified surveillance.

At the disease level, discrimination varied across AML, ALL, and MDS (acute 0.62– 0.64; chronic 0.68–0.79), highest in MDS (**Figure 2 & Figure 4)**, biologically plausible given disease-specific characteristics, conditioning, donor factors, and immune reconstitution^4,5,9^, and suggesting pooled models may mask clinically meaningful subgroup variation.

Beyond discrimination, calibration is the study’s central contribution. Models here are almost always judged by discrimination alone, with the rare calibration figures being single in-sample summaries^15^. Calibration degraded substantially and unevenly externally: models overestimated absolute risk in most cohorts (negative intercept in 9 of 10 evaluable cohort–outcome combinations), while the slope drifted in both directions and by differing magnitudes, so some cohorts needed only an overall reset and others also a correction of spread (**Table 3, Figure 5**). The only exception was P5356 chronic. Crucially, neither direction nor magnitude could be anticipated from training performance or cohort source, so calibration must be re-estimated locally and reported alongside discrimination in every external evaluation^15^. To our knowledge this is the first systematic characterization of multi-cohort calibration drift in GVHD prediction, reporting intercept and slope with full 95% confidence intervals, a message extending well beyond GVHD. Exploratory survival modelling (Supplementary) is intended for future development^13,14,29^.

Feature importance was biologically coherent: acute GVHD was driven mainly by recipient age, graft source, and transplant year, and chronic GVHD by GVHD prophylaxis, conditioning intensity, PBSC use, and HLA compatibility, consistent with established determinants^3,4,5^. The prominence of transplant year for acute GVHD warrants caution, as it may encode era-specific changes in prophylaxis and supportive care rather than a biological effect.

Strengths: the study pairs prediction with SHAP-based explainability, evaluates across six external cohorts rather than a single split, provides the first multi-cohort characterization of calibration drift, archives the full pipeline for exact reproduction, and includes an under-represented Middle Eastern population rarely present in registry-trained models. External cohorts also carried fewer usable features than the training cohort (**Figure 3**), likely contributing to the discrimination drop for both outcomes. But because both models drew on the same features yet only acute collapsed externally **(Figure 4)**, feature coverage alone cannot explain it; the residual acute deficit more plausibly reflects the limited intrinsic signal registry variables carry for acute GVHD
^28,12, 30^.

Several limitations temper interpretation. The five CIBMTR-derived external cohorts share the training data’s predominantly US registry source, with overlapping transplant periods and populations (notably haploidentical PTCy recipients; **Supplementary Document 15**); as releases are de-identified, patient-level overlap cannot be excluded, so these constitute registry-derived rather than fully independent evaluation, although any overlap would inflate, not explain, the observed external degradation. Cohorts differed in data collection and follow-up, and outcomes were taken as adjudicated by each source rather than re-derived, so differences in grading vintage, inter-observer variability, and late-acute or overlap-syndrome presentations may add noise. Biologically relevant variables were often missing (handled natively by CatBoost); binary outcomes ignore death and relapse as competing risks; collinearity between haploidentical donors and PTCy precluded separating their effects; and the Ulschmid acute benchmark is literature-level, with no chronic-GVHD comparator. In UAE+KHCC, acute-GVHD documentation could not be harmonized with registry grading, and chronic-GVHD ascertainment was assumed, but not verified, to follow NIH criteria. Such endpoint mismatch makes calibration fail informatively, reflecting a limit of data transportability rather than of the algorithm; local deployment therefore requires prior endpoint harmonization and per-cohort recalibration, and prospective evaluation is required before any clinical use^28,12,14,31^.

Taken together, these results are a rigorous evaluation of registry-based machine learning for GVHD prediction. The methodological yield is twofold: a multi-cohort external evaluation showing internally modest models can collapse to near chance externally, and a characterization of calibration drift showing that the direction and magnitude of recalibration cannot be anticipated from training performance.

## Conclusion

The central novelty of this work is the first multi-cohort external evaluation of a baseline chronic-GVHD prediction model built entirely from routinely available registry variables, without post-transplant biomarkers or high-dimensional longitudinal data. This multi-cohort evaluation shows it retained limited but above-chance discrimination (AUROC 0.54–0.64) across all external cohorts despite incomplete feature availability and across heterogeneous populations, including an under-represented Middle Eastern cohort; its internal discrimination (AUROC 0.74) exceeded the range typically reported for existing models, albeit as a literature-level comparison. By contrast, acute-GVHD prediction, despite acceptable internal performance, can collapse toward chance externally: using the same feature set, missing data degraded acute more than chronic prediction, and SHAP explanations indicated acute GVHD is driven by different predictors. To our knowledge, this is also the first systematic multi-cohort external evaluation and calibration characterization of GVHD prediction models, establishing that internal performance does not guarantee external utility and that per-cohort recalibration is mandatory, a practical blueprint for validating clinical AI in transplantation. Clinical utility was not assessed and cannot be inferred from these results. The framework is intended for research use only.

## Data Availability

All data produced in the present study are available upon reasonable request to the authors

https://zenodo.org/records/19967153

https://app.gvhdriskcalculator.com/

## Ethics Statement

Analyses of publicly released, de-identified CIBMTR datasets followed CIBMTR data-use policies; secondary methodological use was confirmed with CIBMTR. Use of the UAE and KHCC datasets was approved by the respective institutional review boards (SSMCREC-444; 24KHCC83). This work extends a pilot framework approved by the Abu Dhabi Department of Health.

## Author contributions

N.S.: Conceptualization, Methodology, Software, Data curation, Formal analysis, Investigation, Visualization, Project administration, Writing – original draft, Writing – review & editing. S.H.: Conceptualization, Methodology, Supervision, Writing – review & editing. M.Y.: Methodology, Writing – review & editing. M.A., F.M.A.K., A.R., A.A.Z., F.S., Y.A., D.G., I.A., M.D.: Investigation, Writing – review & editing. H.A.J., H.A.R., K.H.: Resources, Data curation, Writing – review & editing. All authors approved the final version.

## Data availability

The CIBMTR training and external datasets are publicly available from CIBMTR (source studies listed in Supplementary Document 15). The UAE+KHCC data are not publicly available owing to institutional restrictions. The frozen model, code and analysis pipeline are available at https://doi.org/10.5281/zenodo.19967153.

## Declaration of generative AI and AI-assisted technologies

During the preparation of this work, the authors used AI tools including ChatGPT and Claude to assist with improvement of the English language, refinement of scientific writing, simplification and clarification of findings, and enhancement of presentation structure and readability.

All statistical analyses, model outputs, confidence intervals, calibration metrics, ROC curves, SHAP plots, and related visualizations were generated directly by the authors’ machine learning platform and analytical pipeline. The framework is capable of generating standardized analyses and visualizations for additional datasets using the same predefined methodology and output structure.

The authors independently reviewed, verified, and edited all AI-assisted content and take full responsibility for the accuracy, integrity, interpretation, and conclusions of the published work.

## Acknowledgements

The authors gratefully acknowledges Dr. Mheidly Kayane, Dr. Gehad ElGhazali, Dr. Azmat Khan, Dr. Farooq Ahmed Mir, Dr. Riad Al Hasan, Dr. Numan Saeed, and Dr. Muhammad Ridzuan for their contributions to earlier GVHD research projects, including the *GVHD-Intel 1*.*0* study presented at ASH 2025. Their collaboration, clinical insights, and support contributed to several abstracts and publications during the author’s tenure at a previous institution.

The current study was independently developed using the CIBMTR dataset, with data processing, feature engineering, model development, and evaluation performed de novo. The author is grateful for the foundational collaborations that helped advance research in this field.

